# How can we maximise the benefits of smoke-free prisons? Decision analytic model to predict potential impacts on public health

**DOI:** 10.1101/2025.10.30.25339161

**Authors:** Nicola McMeekin, Ashley Brown, Catherine Best, Evangelia Demou, Alastair H. Leyland, Linda Bauld, Nancy Loucks, Jill P. Pell, Sean Semple, Emily J. Tweed, Clair Woods-Brown, Kate Hunt, Kathleen A. Boyd

## Abstract

**Introduction:** Tobacco smoking prevalence remains high in disadvantaged populations such as people in prison. Smokefree prisons protect from smoking harms, however around 90% of people who smoke pre-prison, relapse to smoking shortly after release. If people released from smokefree prisons maintain abstinence this could benefit their health and finances. Knock-on effects of smoking relapse on families could also be avoided. Offering an intervention to reduce relapse to smoking on release has the potential to benefit the health and finances of the released person and their family.

This study assesses potential costs and outcomes for people previously imprisoned and their families, of smokefree prison policy and a hypothetical smoking cessation intervention.

**Methods:** A Markov model was developed for three populations: people previously in prison, partners, and children. Model states are based on smoking/vaping status. Four scenarios were modelled depending on whether vaping is permitted in prison and whether a hypothetical intervention was applied. The analysis was from healthcare payer and personal perspectives, over a lifetime. Costs include healthcare, personal (vaping and smoking) and the intervention. Outcomes include quality adjusted life-years. Several plausible scenarios were conducted due to uncertainty in parameters.

**Results:** Offering an intervention was beneficial for both vaping scenarios. For the person released from prison, results indicated that specific scenarios without vaping permitted in prison were more cost-effective (less costly and more beneficial) than with vaping permitted. However, sensitivity analyses highlighted the significant impact of vaping prevalence and relapse rates on outcomes, emphasising uncertainty in some parameters.

For both partner and child(ren), costs were higher and quality adjusted life-years lower for those living with a released prisoner who smokes compared to one who vapes or does not smoke/vape.

## Interpretation

There is a need for robust evidence on relapse to smoking on release and the long-term effects of vaping. Targeted support for smoking cessation interventions to improve health outcomes for people previously in prison and their families, can ultimately contribute to broader public health improvements and reduce health inequalities. Study findings enhance understanding of the potential cost-effectiveness of smokefree prison policy and can inform decisions about how value could be maximised.

## Introduction

Whilst smoking prevalence overall is decreasing, smoking rates and associated smoking related illness, and exposures to second-hand smoke (SHS), remain relatively high in disadvantaged populations(1, 2), contributing to inequalities in health. Most people in prison are from lower socioeconomic backgrounds, and historically smoking levels in prisons have been high(3, 4). In Scotland, for example, smoking prevalence among the prison population was 68% in 2017(5). A smokefree prison policy was introduced in Scotland in November 2018: no-one (including staff, people in prison and visitors) can smoke in indoor or outdoor areas of all prisons. Evidence suggests that the smokefree prison policy in Scotland effectively eliminated tobacco use and exposure to SHS in prisons(6). Research shows that total smokefree prison policies, like Scotland’s policy, have reduced cardiovascular disease risk among people in prison who smoked prior to imprisonment(7, 8), and are associated with a 9% reduction in smoking related deaths in prison(9).

As part of the transition to smokefree prisons in Scotland, rules were changed to allow people in prison to vape, with 60% reporting vaping in 2019 in Scottish prisons(10). The UK National Institute for Health and Care Excellence (NICE) supports vaping for smoking cessation(11), as evidence to-date suggests vaping is likely to be less harmful than smoking. There is currently insufficient evidence to quantify long-term risks of vaping(12–14).

If people who smoked prior to imprisonment in a smokefree prison were able to maintain abstinence post-release, there could be considerable health and financial benefits for individuals, and reduced pressures on healthcare services and spending. Studies of prison systems which are smokefree and *do not* allow vaping, consistently find a rapid and high return to smoking post-release(15). This highlights a lost opportunity to address a leading cause of inequality in morbidity and mortality; evidence shows that disadvantaged groups are less likely to successfully quit smoking despite being motivated to quit(16). To date, there is no evidence on smoking relapse rates among people released from smokefree prison systems which *do* allow vaping(15). On one hand, switching from smoking to vaping in prison could decrease the likelihood of post-release smoking relapse by providing a potentially cheaper alternative. On the other hand, people who vape in prison may relapse to smoking at a similar or even higher rate due to maintenance of nicotine dependence. Dual vaping and smoking might also increase post-release with little if any benefit to health.

Secondary negative impacts of smoking on family members could also be averted if those who smoked prior to imprisonment in a smokefree prison remained abstinent post-release. In Scotland, over 20,000 children are affected by parental imprisonment every year(17). Having a smoking parent increases the likelihood of children being exposed to SHS in homes and experiencing smoking-related diseases(18, 19). Children living with people who smoke are also more likely to start smoking themselves(20). Smoking disproportionately impacts the finances of families living on low incomes and exacerbates child poverty(21, 22). Imprisonment can place considerable (additional) financial strain on families. Respondents to a recent survey reported spending a third of their monthly income on a family member in prison, and around a half of their monthly income to support a family member after release(23). Reducing or eliminating smoking costs could therefore make a substantial difference to the lives of families affected by imprisonment.

The multi-phase, multi-method Tobacco in Prisons Study (TIPS) assessed the impact of introducing smokefree prison policy in Scotland(24). An economic evaluation found that introducing the policy was cost-effective for people in prison and staff over a lifetime(25), although limited data were available to inform some estimates (e.g. post-release relapse rates). We originally planned to try to address these evidence gaps in a follow-up study, update the lifetime model for people in prison and, for the first time, model the costs and outcomes of smokefree prison policies for families affected by imprisonment. However, the effects of the COVID-19 pandemic in prisons, combined with other factors, prevented this. The current study therefore assesses the potential costs and outcomes of smokefree prison policy for people previously imprisoned and their families under a range of plausible scenarios. Findings from the study can increase understanding of the potential cost-effectiveness of smokefree prison policy and inform decisions about how value could be maximised.

## Methods

### Overview

A modelling approach was taken. The populations modelled were people recently released from a smokefree prison (hereafter referred to as ‘released person/people’), their partners and children (from age 15) in a Scottish household setting, with costs and outcomes estimated over a lifetime.

Costs and outcomes were included from both the healthcare payer (public purse) and personal (cost to individuals) perspectives. Due to limitations imposed by the pandemic and in the global evidence base, as noted above, we have included a hypothetical intervention to support smoking cessation and prevent relapse to smoking post-release (hereafter referred to as a ‘smoking cessation intervention’) in the model. A discount rate of 3.5% was applied to costs and quality adjusted life-years (QALYs) in line with current recommendations from NICE(26).

Methods and results are presented in line with established guidance(27).

### Comparators

We modelled four scenarios depending on a) whether vaping is permitted in smokefree prisons and b) whether a smoking cessation intervention is offered around the time of release (Table 1).

**Table 1.**
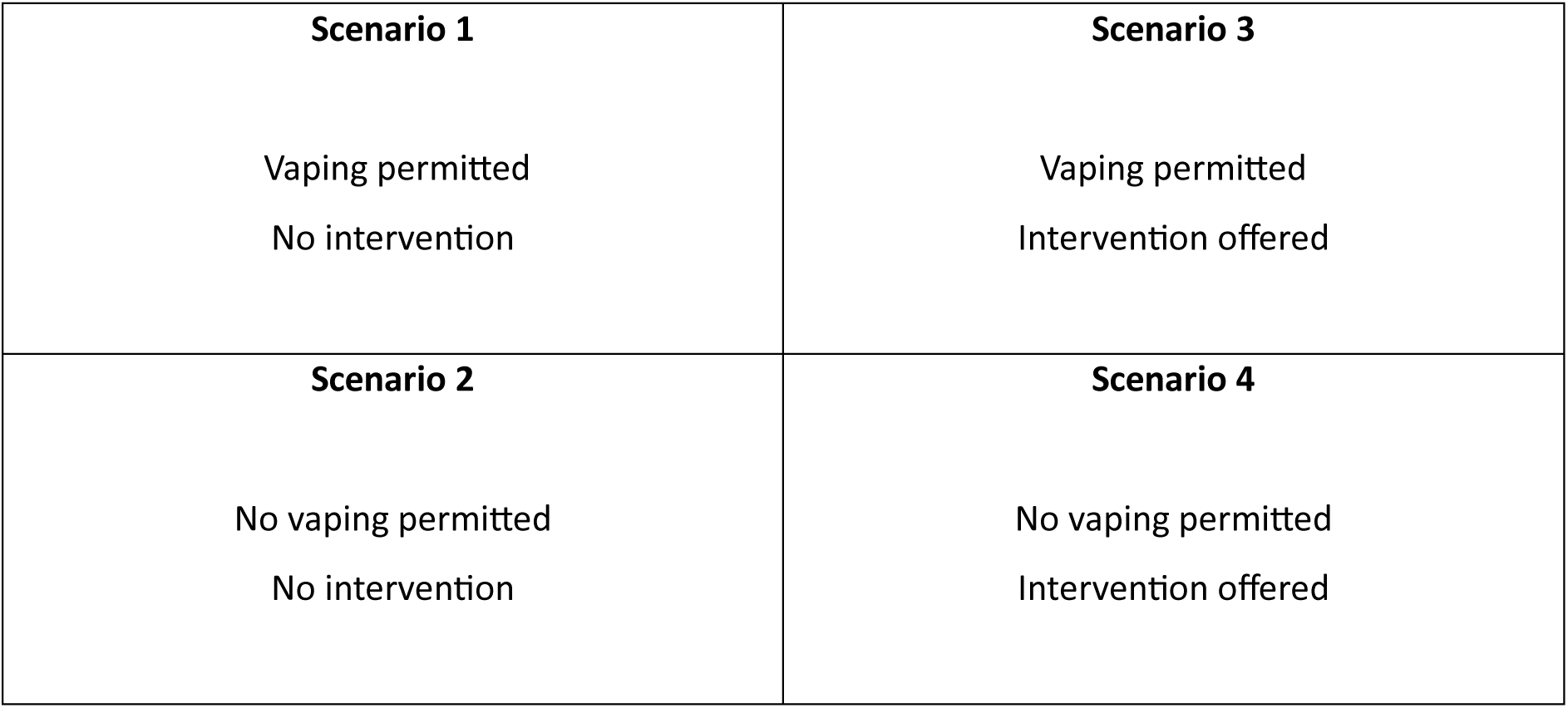
Model scenarios.

To evaluate cost-effectiveness we compared: i) vaping permitted in smokefree prisons v no vaping permitted in smokefree prison (1 v 2); ii) no intervention vs intervention offered (vaping permitted) (1 v 3), and iii) no intervention vs intervention offered (no vaping permitted) (2 v 4).

### Model structure

Figure 1 illustrates the structure and seeding of three sets of models: for released people from a prison which (a) allows vaping or (b) does not allow vaping; and (c) for their partners/children. Due to the lack of evidence on the proportion of released people who return to a family home with a partner and/or children, we have not included a link between the released people model and the partner/child models; results for partner/child models are presented separately.

**Figure 1.**
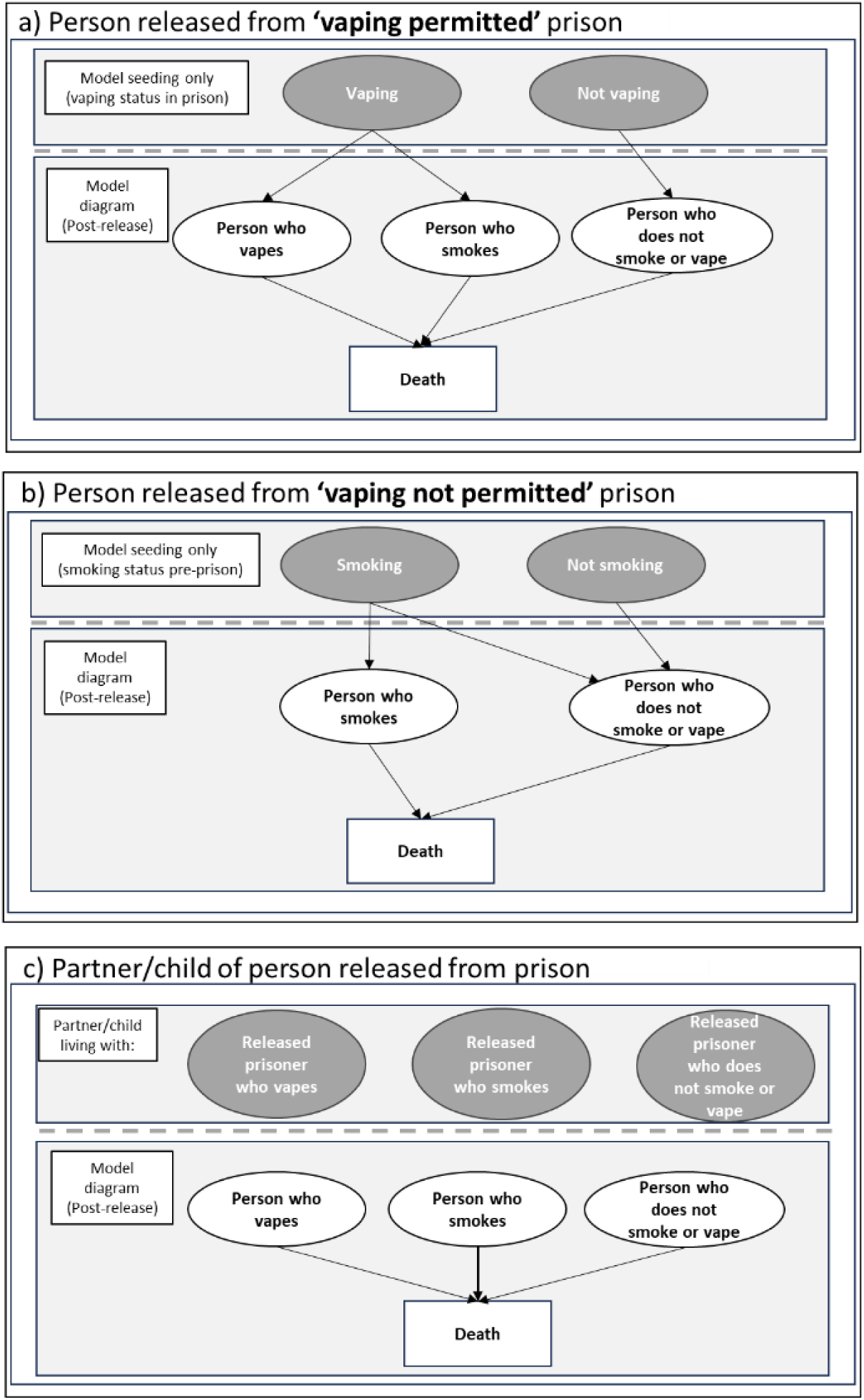
Model diagram

The decision analytic models were built with four states, three relating to smoking/vaping (nicotine) states and one Death state. The three nicotine states were: ‘Person who vapes’; ‘Person who smokes (tobacco)’, and ‘Person who neither smokes nor vapes’ (hereafter described as ‘nicotine-free’). The model is run for 60 annual cycles for released people and partners, and for 100 years for children. For each cycle, model participants remain in their nicotine state or transition to the ‘Death’ state.

Participants accumulate costs and health utilities as they move through the model until they join the ‘Death’ state.

All model parameters are presented in Supplementary Material Table S1.

### Smoking/vaping status

Smoking/vaping status for released people in the first model cycle is dependent on: a) vaping status in ‘vaping-permitted’ prisons (calculated by CB using anonymised SPS ‘canteen’ purchasing data), or b) smoking status prior to imprisonment in ‘no vaping permitted’ prisons(5).

In the ‘vaping-permitted’ scenarios (a), we assume people who vape in prison will either smoke or vape post-release, and people who do not vape in prison will remain nicotine-free. As a recent scoping review found no evidence on smoking/vaping rates following release from a smokefree prison which permits vaping(15), we have applied evidence from smokefree prisons where vaping is not permitted: basecase is 92% of people who vape in prison will smoke on release and 8% will vape(28). The same evidence is applied in the ‘vaping not permitted’ scenarios (b); we assume for the basecase that 92% of people held in prison who smoked prior to imprisonment relapse on release, the remaining 8% will be nicotine-free(28).

Smoking status for partners is taken from Scottish Health Survey 2022, using rates for women living in the most deprived neighbourhoods(29).

We assume that child uptake of smoking/vaping is dependent on the vaping or smoking status of the released person. An annual probability of smoking uptake is applied to children if the parent does not smoke(30). Children who live with a released person who smokes or vapes have an additional risk of smoking/vaping uptake applied(31).

### Morbidity

Smoking related disease (SRD) prevalence (chronic obstructive pulmonary disease (COPD), coronary heart disease (CHD), lung cancer and stroke) was applied to all models(32), dependent on sex, age and smoking status. General population SRD prevalence was applied for nicotine-free people and people who vape(30). Additional age and sex specific relative risks were applied to model participants who currently or used to smoke(32).

There is emerging mixed evidence on vaping-related co-morbidities; after consultation with a chest physician, we applied acute bronchitis incidence to people who vape, which we consider to be a conservative approach(33, 34).

To account for SHS exposure, an increase in risk for CHD and lung cancer was applied to nicotine-free partners who live with a released person who smokes(35).

### Mortality

Mortality rates were applied to all models, dependent on sex, age and smoking status. Scottish general population mortality rates were applied to nicotine-free people and people who vape(36). Increased risk of mortality for people who currently smoke or used to smoke was applied(37), incorporating sex and age specific smoking prevalence for Scottish population(29).

A further adjustment was made for the known increased risk of mortality for released people(38–40). Finally, current evidence suggests that 1% of global mortality can be attributed to SHS exposure; we incorporated this into our model for partners living with released people who smoke(35).

### Smoking cessation intervention

We assume that a hypothetical intervention to promote smoking cessation and prevent relapse to smoking (see below) is available to people who smoked prior to imprisonment in the ‘no vaping permitted’ scenarios, and to people who vape in ‘vaping permitted’ prisons. The effectiveness of the intervention is applied in the model in the first year after release. To model intervention impact we applied the costs and effectiveness of a medium intensity smoking cessation intervention reported in NICE smoking guidelines for the general population(11). We explored the impact of low and high intensity interventions on the results in sensitivity analyses. Low intensity consists of brief advice and self-help materials, medium intensity adds nicotine replacement therapy, and high intensity adds nicotine replacement therapy and specialist smoking cessation services(41).

We assume that the proportion of the model cohort engaging in the intervention is equal to the number of people in prison reporting a desire to quit smoking in the 16^th^ SPS Prisoner Survey(10).

### Costs

Resource use comprises healthcare (SRD treatment and harms from vaping), personal (smoking and vaping products), and intervention. Unit costs were applied to resources, sourced from literature and published reports: bronchitis(42); SRD(43); personal nicotine products(44), and intervention(11). Unit costs are measured in GBP for 2021/22. Where relevant, costs were inflated using NHS cost inflation index (NHSCII)(45).

### Outcomes

Outcomes include quality adjusted life-years (QALYs), life-years (life expectancy) and prevalence of smoking/vaping/nicotine-free. The QALY is a combination of quality-of-life (health utilities) and length of life (life-years). Age-specific health utilities were taken from the literature for the general population to represent nicotine-free people and people who used to vape(46). A reduction in health utilities (disutility) was applied for people who smoke, vape, who used to smoke, and partners exposed to SHS. Disutilities were sourced from literature for people who smoke and used to smoke(47) and people who vape(48), and from the TIPs results for the impact of SHS(25). Disutilities for SRD were also applied(49).

Prevalence is extracted from the first cycle immediately after release.

### Further model assumptions

SHS exposure is only applied to partners; we assume that children are only exposed to SHS for a short period before they no longer live with the released person (starting in the model at 15 years old).

For morbidity and mortality risks, released people are considered to be male (Scottish Prisoner Survey reported 93% are male(10)), partners female, and children 50% male and 50% female.

The cost of bronchitis is based on reported spilt between inpatient and non-hospitalised treatment for COPD exacerbations, applying the cost of a mild to moderate exacerbation(42).

### Analysis

A cost-utility analysis was conducted for released people using the three comparisons described above. Mean costs, QALYs and life-years are presented with 95% confidence intervals (95%CI).

Incremental results are used to estimate incremental cost-effective ratios (ICERs). Incremental net monetary benefit (iNMB) is calculated using a willingness-to-pay threshold of £20,000; iNMB is straightforward to interpret with results above zero indicative of cost-effectiveness.

Partner and child results (mean costs, QALYs and life-years with 95%CI) are presented for three household scenarios where the partner/child lives with a released person who is either nicotine-free, smokes, or vapes.

Uncertainty in the model parameters was assessed with probabilistic sensitivity analysis using best practice methods(50). Results were used to estimate 95%CI around costs and QALYs. Results are illustrated in a cost-effectiveness plane.

### Sensitivity Analyses

To understand the potential of smokefree policies and due to uncertainty in the parameters, several sensitivity analyses were conducted, in addition to the previously described basecase, varying: vaping prevalence in prison; relapse to smoking on release; harms from vaping; engagement with intervention; intervention success (e.g. successful quits) rate; and uptake of smoking in children with parents who do not smoke or vape. Results are illustrated in a tornado diagram (released person) and table (released person and child) using iNMB.

## Results

### Released person

For basecase the largest proportion of costs in all scenarios were for personal (smoking/vaping) costs (around 78% of total costs), with healthcare costs making up just over 20% of total costs.

Intervention costs were around £40 (<1%). Total costs ranged from £26,880 (95%CI £22,392, £32,169) for Scenario 4, to £31,347 (95%CI £26,451, £37,271) for Scenario 1 (Table 2).

**Table 2.** Results – Released person.

|  | No intervention |  |  |  | Intervention |  |  |  |
| --- | --- | --- | --- | --- | --- | --- | --- | --- |
|  | Vaping permitted in smokefree prison (Scenario 1) |  | No vaping permitted in smokefree prison (Scenario 2) |  | Vaping permitted in smokefree prison (Scenario 3) |  | No vaping permitted in smokefree prison (Scenario 4) |  |
|  | Mean | 95% CI | Mean | 95% CI | Mean | 95% CI | Mean | 95% CI |
| **Costs** |  |  |  |  |  |  |  |  |
| Healthcare | £ 6,673 | £5,930 to £7,533 | £6,137 | £5,467 to £6,957 | £6,393 | £5,681 to £7,220 | £6,080 | £5,423 to £6,877 |
| Personal | £ 24,674 | £19,870 to £30,175 | £21,446 | £16,972 to £26,449 | £23,889 | £19,238 to £29,215 | £20,764 | £16,432 to £25,608 |
| Intervention | - | £0 to £0 | - | £0 to £0 | £43 | £43 to £43 | £36 | £33 to £37 |
| **Total** | £ 31,347 | £26,451 to £37,271 | £27,584 | £22,953 to £33,039 | £30,325 | £25,585 to £36,054 | £26,880 | £22,392 to £32,169 |
| **Life-years** | 17.20 | 16.89 to 17.48 | 17.39 | 17.11 to 17.66 | 17.23 | 16.94 to 17.51 | 17.42 | 17.16 to 17.68 |
| **QALYs** | 13.54 | 13.22 to 13.81 | 13.79 | 13.50 to 14.06 | 13.58 | 13.28 to 13.85 | 13.82 | 13.54 to 14.06 |
| **Prevalence** |  |  |  |  |  |  |  |  |
| Non-nicotine | 24% |  | 37% |  | 24% |  | 37% |  |
| Person who smokes | 70% |  | 63% |  | 68% |  | 61% |  |
| Person who used to smoke | 0% |  | 0% |  | 2% |  | 2% |  |
| Person who vapes | 6% |  | 0% |  | 6% |  | 0% |  |
| Person who used to vape | 0% |  | 0% |  | 0% |  | 0% |  |

Cost-utility analysis – comparing Scenarios
|  | Permitting vaping v. not permitting vaping with no intervention in smokefree prison (Difference 1) |  | Vaping permitted in smokefree prison (Difference 2) |  | No vaping permitted in smokefree prison (Difference 3) |  |
| --- | --- | --- | --- | --- | --- | --- |
|  | Vaping permitted (Scenario 1) | No vaping permitted (Scenario 2) | No intervention (Scenario 1) | Intervention (Scenario 3) | No intervention (Scenario 2) | Intervention (Scenario 4) |
| **Total costs** | £31,347 (95%CI £26,451 to £37,271) | £27,584 (95% CI £22,953 to £33,039) | £31,347 (95%CI £26,451 to £37,271) | £30,325 (95%CI £25,585 to £36,054) | £27,584 (95% CI £22,953 to £33,039) | £26,880 (95% CI £22,392 to £32,169) |
| **Total QALYs** | 13.54 (95% CI 13.22 to 13.81) | 13.79 (95% CI 13.50 to 14.06) | 13.54 (95% CI 13.22 to 13.81) | 13.58 (95% CI 13.28 to 13.85) | 13.79 (95% CI 13.50 to 14.06) | 13.82 (95% CI 13.54 to 14.06) |
| **Incremental (costs and QALYs)** | £3,764 (95% CI £2,715 to £5,125) | -0.252 (95% CI -0.355 to -0.17) | £1,022 (95% CI £866 to £1,209) | -0.044 (95% CI -0.056 to -0.033) | £704 (95% CI £561 to 871) | -0.037 (95% CI -0.126 to 0.045) |
| **Cost per QALY** | Dominated (more costly and less effective) |  | Dominated (more costly and less effective) |  | Dominated (more costly and less effective) |  |
| **iNMB** | -£8,798 |  | -£1,911 |  | -£1,437 |  |

iNMB–incremental net monetary benefits; QALY–quality adjusted life-year

There was little variation in life-years and QALYs between scenarios, which ranged from 17.20 (95%CI 16.89, 17.48) (Scenario 1) to 17.42 (95%CI 17.16, 17.68) (Scenario 4), and 13.54 (95%CI 13.22, 13.81)

(Scenario 1) to 13.82 (95%CI 13.54, 14.06) (Scenario 4) respectively.

The least costly and most effective scenario was Scenario 4 (no vaping permitted, intervention offered), the most costly and least effective was Scenario 1 (vaping permitted, no intervention offered).

The prevalence of post-release smoking is highest in Scenario 1 (70%) (no intervention) and lowest in Scenario 4 (61%) (intervention offered). Vaping is 6% in Scenarios 1 and 3.

The cost-utility analysis results show that in Difference 1 the scenario permitting vaping is more costly (£3,764 (95%CI £2,715, £5,125)) and less effective (QALYs-0.252 (95%CI-0.355,-0.170)) than not permitting vaping. In Difference 2, when vaping is permitted, not offering an intervention is more costly (£1,022 (95%CI £866, £1,209)) and less effective (QALYs-0.044 (95%CI-0.056,-0.033)) than offering an intervention. In Difference 3 when vaping is not permitted, not offering an intervention is more costly (£704 (95%CI £561, £871)) and less effective (QALYs-0.037 (95%CI-0.126, 0.045)) than offering an intervention.

These results are reflected in iNMB results with no results above £0; suggesting that with no intervention not permitting vaping is more cost-effective than permitting vaping, and offering an intervention for both vaping permitted and vaping not permitted is more cost-effective than not offering an intervention.

The cost-effectiveness plane (Figure 2) illustrates the above results visually, the incremental costs and QALYs are plotted in four quadrants which show which comparators are more/less costly and effective. Differences 1 (vaping permitted v no vaping permitted with no intervention) and 2 (no intervention v intervention in vaping permitted prison) fall entirely in the more expensive and less effective area of the graph. Difference 3 (no intervention v intervention in no vaping permitted prison) results show that whilst Scenario 2 is consistently more expensive than Scenario 4, there is some uncertainty in the QALY results which straddle the horizontal axis, reflected in the 95%CI presented above.

**Figure 2.**
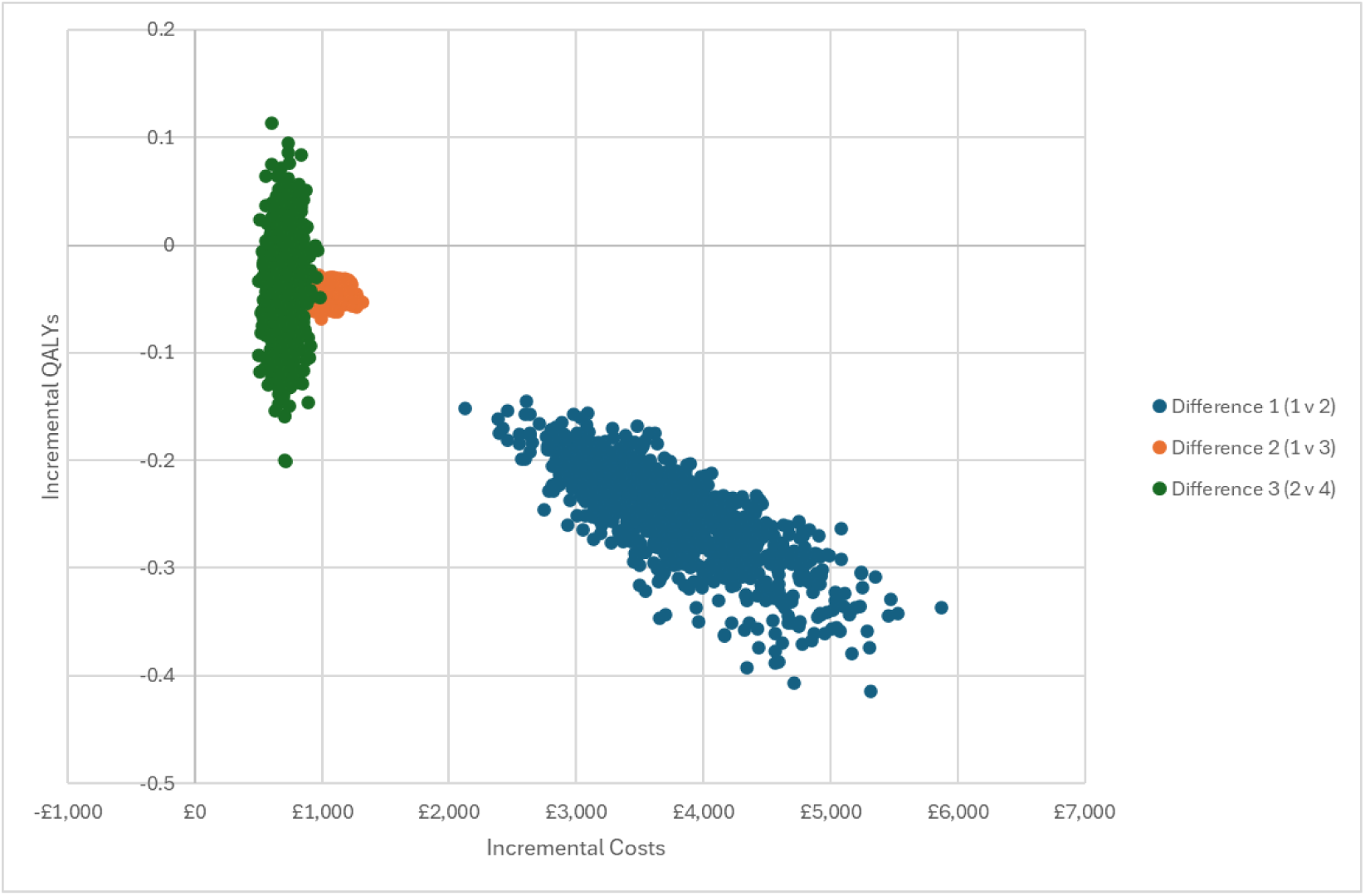
Cost-effectiveness plane – Released person

**Figure 3.**
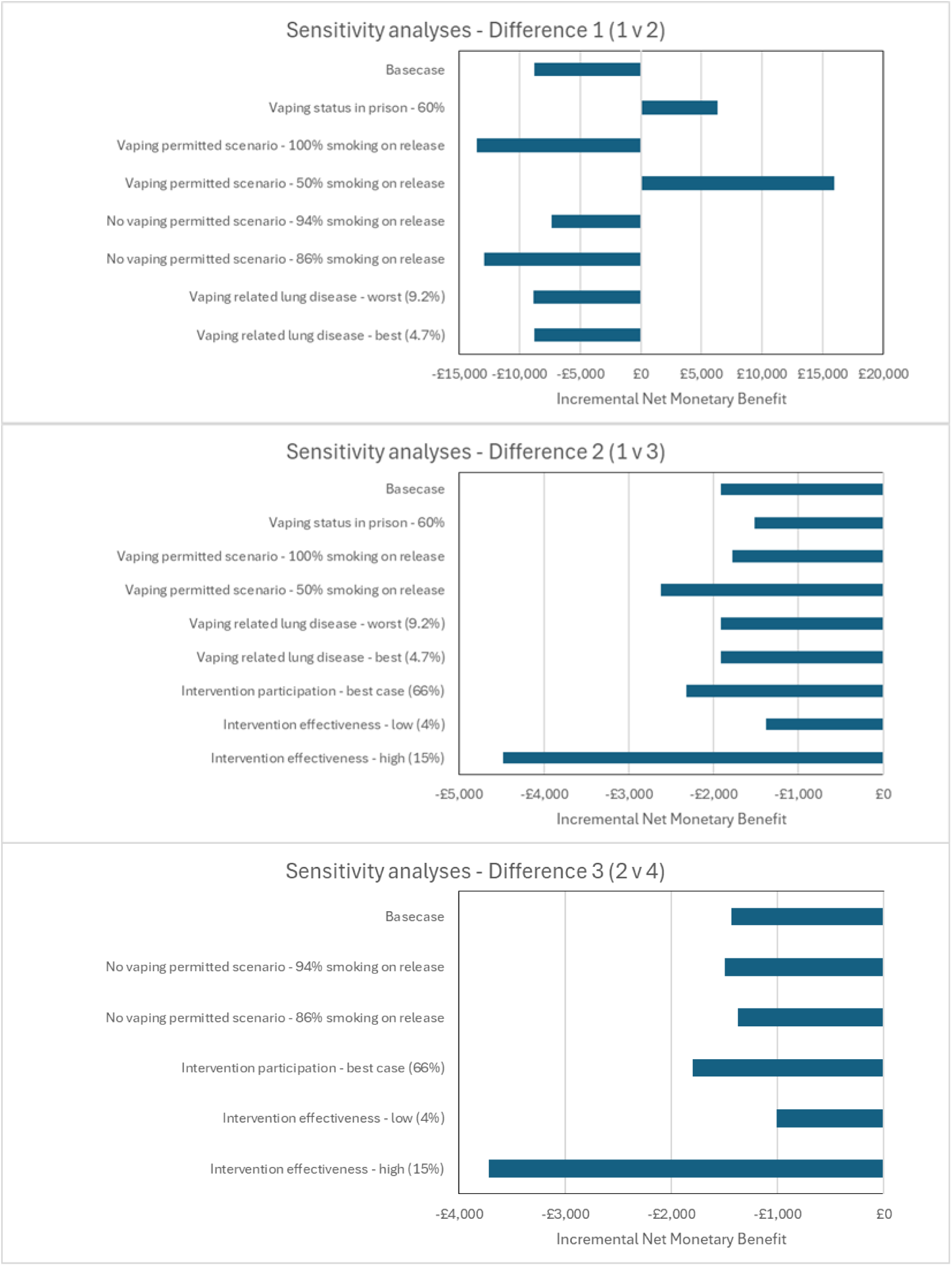
Sensitivity analyses results

### Partner and child

For basecase the highest costs for partners were personal costs (smoke and vape products) for all three household living circumstances (Table 3), around two thirds of total costs. Healthcare costs were around one third of total costs and slightly higher in partners living with a released person who smokes due to SHS exposure, (£5,547 (95%CI £5,053, £6,090)) compared to £5,295 (95%CI £4,821, £5,809)) where the released person is nicotine-free. There is very little difference in life-years and QALYs between different living circumstances: 22.15 v. 22.16 and 17.54 v. 17.82 respectively.

**Table 3.** Results - Parent and child.

|  | Living with nicotine-free person released from prison |  | Living with person released from prison who smokes |  | Living with person released from prison who vapes |  |
| --- | --- | --- | --- | --- | --- | --- |
|  | Mean | 95% CI | Mean | 95% CI | Mean | 95% CI |
| **<u>Partner</u>** |  |  |  |  |  |  |
| **<u>Costs</u>** |  |  |  |  |  |  |
| Healthcare | £5,295 | £4,841 to £5,809 | £5,547 | £5,053 to £6,090 | £5,295 | £4,813 to £5,785 |
| Personal | £10,617 | £8,157 to £13,610 | £10,617 | £8,157 to £13,610 | £10,617 | £8,157 to £13,610 |
| Intervention | - |  | - |  | - |  |
| Total | £15,912 | £13,212 to £19,018 | £16,164 | £13,449 to £19,280 | £15,912 | £13,212to £19,018 |
| Life-years | 22.16 | 22.06 to 22.25 | 22.15 | 22.04 to 22.24 | 22.16 | 22.06 to 22.25 |
| QALYs | 17.82 | 17.61 to 18.01 | 17.54 | 17.36 to 17.70 | 17.82 | 17.61 to 18.01 |
| **<u>Prevalence</u>** |  |  |  |  |  |  |
| Non-nicotine | 63% |  | 0% |  | 63% |  |
| Non-nicotine (exposed to SHS) | 0% |  | 63% |  | 0% |  |
| Person who smokes | 21% |  | 21% |  | 21% |  |
| Person who used to smoke | 0% |  | 0% |  | 0% |  |
| Person who vapes | 10% |  | 10% |  | 10% |  |
| Person who used to vape | 0% |  | 0% |  | 0% |  |
| **<u>Child</u>** |  |  |  |  |  |  |

| **Costs** |  |  |  |  |  |  |
| --- | --- | --- | --- | --- | --- | --- |
| Healthcare | £2,800 | £2,563 to<br>£3,067 | £3,596 | £3,264 to<br>£3,966 | £3,282 | £2,989 to<br>£3,607 |
| Personal | £10,312 | £8,542 to<br>£12,495 | £24,875 | £20,472<br>to £30,386 | £19,022 | £15,671 to<br>£23,198 |
| Intervention |  |  |  |  |  |  |
| Total | £13,113 | £11,268 to<br>£15,300 | £28,471 | £24,009 to<br>£34,046 | £22,304 | £18,876 to<br>£26,552 |
| **Life-years** | 25.69 | 25.66 to<br>25.70 | 25.49 | 25.42 to<br>25.54 | 25.56 | 25.51 to<br>25.60 |
| **QALYs** | 22.02 | 21.89 to<br>22.13 | 21.41 | 21.18 to<br>21.62 | 21.66 | 21.47 to<br>21.83 |
| **Prevalence** |  |  |  |  |  |  |
| Non-nicotine | 70% |  | 33% |  | 49% |  |
| Non-nicotine (exposed<br>to SHS) | 0% |  | 0% |  | 0% |  |
| Person who smokes | 15% |  | 39% |  | 30% |  |
| Person who used to<br>smoke | 0% |  | 0% |  | 0% |  |
| Person who vapes | 15% |  | 27% |  | 22% |  |
| Person who used to vape | 0% |  | 0% |  | 0% |  |

*CI—confidence interval; SHS—second-hand smoker; QALY—quality adjusted life-year*

The highest costs for children (from age 15) were personal costs for all living circumstances, ranging from £10,312 (95%CI £8,542, £12,495) for living with a nicotine-free released person to £24,875 (95%CI £20,472, £30,386) for living with a released person who smokes. Healthcare costs varied less than personal costs, ranging from £2,800 (95%CI £2,563, £3,067) for living with a nicotine-free released person to £3,596 (95%CI £3,264, £3,966) living with a released person who smokes. For outcomes life-years ranged from 25.49 (95%CI 25.42, 25.54) to 25.69 (95%CI 25.66, 25.70) and QALYs from 21.41 (95%CI 21.18, 21.62) to 22.02 (95%CI 21.89, 22.13). The prevalence of children who smoke [or vape] ranges from 15% [15%] for children who live with a nicotine-free released person to 39% [27%] for children who live with a released person who smokes.

### Sensitivity analyses

Results are presented in **Error! Reference source not found.** and Supplementary Material **Error! Reference source not found.** for released people. In Difference 1 (comparing vaping permitted v no vaping permitted smokefree prisons) permitting vaping in smokefree prisons becomes the more cost-effective scenario if the vaping rate is lowered to 60% or the smoking relapse rate is lowered to 50% (reversing basecase results). In contrast, the two parameter changes which result in biggest negative impact on cost-effectiveness are assuming a) 100% of people who vape in prison (vaping permitted) returning to smoking on release and b) 86% of people who smoked prior to entering prison (no vaping permitted) relapse to smoking on release.

In Difference 2 (comparing vaping permitted policies with/without an intervention) the two parameter changes with the biggest impact on results (making the hypothetical intervention less costly and more effective) are assuming a) 50% of people who vape in prison relapse to smoking post-release, and b) the intervention is highly effective. Both result in a lower iNMB than in basecase analysis.

In Difference 3 (comparing vaping not permitted policies with/without an intervention) the two parameters with the biggest impact on results (making the intervention less costly and more effective) are: a) increasing intervention engagement to 66% and b) increasing the effectiveness of the intervention. Both result in a lower iNMB than in basecase analysis.

For the sensitivity analysis impacting the children’s model results (Supplementary Material Table S3), increasing the uptake of smoking/vaping for children living with a parent who smokes from 15.2% basecase to 22.8% worst case, results in total costs increasing and total QALYs decreasing.

Conversely, when the uptake is decreased to 7.6% best case, total costs are lower and QALYs are higher. In the worst case analysis, all children either took up smoking (59%) or vaping (41%).

## Discussion

For basecase results, permitting vaping in a smokefree prison resulted in higher costs and less QALYs compared to no vaping permitted. While we have assumed 92% of people who vape in prison or were smokers prior to imprisonment, will return to smoking post-release in both the vaping permitted and no vaping permitted scenarios respectively, different seeding assumptions for the two scenarios mean that we expect higher numbers of people released from a vaping-permitted prison to return to smoking, compared to a no vaping permitted prison. However, the results are reversed in the sensitivity analysis which assumes a lower vaping prevalence in prison (60% vs 76% basecase), highlighting the impact of uncertainty about the values for vaping prevalence in prisons (and smoking prevalence pre-imprisonment in the no vaping permitted scenario) on study results. The sensitivity analysis assuming 100% of people who vape in prison relapse to tobacco smoking on release (92% basecase), resulted in higher incremental costs and QALYs, whereas when this parameter was lowered to 50% smoking on release, the vaping permitted policy became more cost saving and more effective. Again, our study highlights the impact of uncertainty in evidence on the rates of post-release smoking relapse among people who vaped in prison. Offering a smoking cessation intervention to people with experience of imprisonment was cost saving (for personal and healthcare costs) and more effective in both the vaping permitted and no vaping permitted scenarios. This finding did not change in any of the sensitivity analyses. Additionally, we found that offering a more (rather than less) effective intervention had the biggest positive impact on results from all sensitivity analyses. In both the partner and child models, the most costly scenario was living with a released person who smokes. In all models there was little variation in life-year and QALY outcomes between scenarios, however sensitivity analyses results show that applying higher intervention effectiveness improves these outcomes, reflecting the benefits of an effective intervention.

### Limitations

The biggest limitation is lack of evidence in this field: we went to considerable effort to source the most robust evidence available, but lack of reliable evidence remains a major constraint. The most substantial gap in the evidence that we found was smoking status on release from smokefree prisons permitting vaping; the best evidence we had for relapse to smoking on release was from a ‘no vaping permitted’ smokefree prison system in Australia which we also applied to our ‘vaping permitted’ scenarios in the absence of any other evidence(15, 28).

There is limited evidence to date on the extent of health harms from vaping. Whilst UK NICE recommends the use of e-cigarettes for smoking cessation(11), there is emerging evidence that vaping(12–14) and exposure to second-hand aerosols from e-cigarettes(51, 52) may be harmful to health. We used a conservative approach and simply considered the increased risks around acute bronchitis from vaping. It is noteworthy that future evidence on vaping-related harms from general population studies may not be applicable to released people; evidence to date suggests that people in prison have different ways of using vapes, including heavy use and consumption of illicit substances(53, 54).

Unfortunately, due to the impact of COVID-19 on the prison system, we were unable to develop a bespoke intervention to support released prisoners to remain smoking abstinent, so had to base parameters on a hypothetical intervention. There is a small amount of intervention effectiveness evidence (for post-smokefree prison smoking abstinence) from Australia(55–57), but this was limited to short time periods and from small studies. Therefore we based our parameters on a NICE model with low, medium and high effectiveness and related costs(11) developed for a general population of smokers. We assume that our intervention is offered to people who do not want to relapse to smoking on release, either to people who are about to be released and/or who have recently been released.

There is also a lack of evidence for living status on release; we were unable to source evidence on how many people would live with partners/parents/a friend/alone or homeless/temporary housing when released from custody. As a solution to this problem, we modelled partners and children separately, and readers can use results that are relevant to their needs.

Scarce evidence for uptake of smoking or vaping in children living with a parent who smokes or vapes resulted in us using evidence for ‘ever vapers’ and ‘ever smokers’(31), and we are aware that patterns of vaping in children and young people have been changing quickly in recent years.

As in all models we made simplifying assumptions. For example, we did not take account of reimprisonment, although we know that many released people may return to prison at a later date(58). This makes our models conservative, as reimprisonment removes the ability to smoke tobacco during the prison stay and any intervention offered to reduce relapse to smoking would potentially be more beneficial if people released from prison are exposed to it multiple times. We did not include dual users of tobacco and vapes; there was a lack of evidence for specific harms in this population and no evidence on how many released people would be dual users. We assumed that partner smoking status is not influenced by the smoking status of the released person as, again, there is no evidence in this area. We also applied partner smoking status based on women living in the most deprived areas. We assumed that released people are men and their partners are women, but are aware that this is an oversimplification. Lastly, our assumption that children start in the model at age 15 likely underestimates cost savings and benefits for this cohort had the model started at a younger age for children.

People in prison are known to have a high mortality rate immediately after release(38), making it hard to separate out complexities in this population to inform a model: our simplifying assumptions are unable to fully assess the relationships between and within these complexities. Additionally, the prison population is more deprived and likely to experience higher mortality than the general population.

### Implications

Smoking continues to be a public health priority; populations with high smoking prevalence are at high risk of smoking related morbidity and mortality. Our results emphasise the importance of providing support to remain smokefree post-release which may include support for those who wish to cut down or quit vaping whilst imprisoned in a smokefree prison where vaping is allowed. There are potential cost-savings, both to individuals and healthcare providers, resulting from offering an intervention, and not offering an intervention is likely to result in harms to released people and to their families. This research found that offering support to remain smoking abstinent post-prison is beneficial, however there are many challenges to implementing such an intervention in this population, for example in terms of resources, person in prison/family appetite, reach for those with unstable living conditions and general logistics.

Current government policy is for the UK to become smokefree by 2030(59), however the Khan Report states that, for the most deprived groups in society, this target is only likely to be met in 2044, leaving residual populations with high smoking prevalence. More resource is needed to support these populations to become smokefree(60). By improving the health of disadvantaged groups (such as people with drug and alcohol dependence, people who are homeless and people in prison/released), the health of the general population will be improved and inequalities will decrease(61). Rates of ill health are much higher in these disadvantaged groups, and although they are small proportion of the population, improving their health will improve the health of the general population.

### Future research

Our previous work recommended further exploring: relapse to smoking post-release; spillover effects of smoking status on households post-release, and impacts of e-cigarette harms in the prison population(25). This study has partially addressed these recommendations. A scoping review, conducted as part of the TIPS2 study, has provided a foundation for evidence on relapse rates to smoking after leaving a smokefree prison(15). This modelling study provides initial evidence on the knock-on effects of the smoking status of released people on household members and on the post-prison period in general. It also provides information on the potential benefits of offering an intervention which could help to develop future interventions and policies. However, more research is needed in these areas, and in particular, more research is needed in identifying vaping harms in the general population and in the prison population and developing effective interventions for this population remains a priority.

## Conclusion

Offering an intervention to support released people to remain smoking abstinent has the potential to be a cost-effective approach, in both vaping scenarios. In a scenario where vaping prevalence is 76% and permitting vaping in prison does not change risk of post-release smoking relapse, permitting vaping is not cost-effective compared to not permitting vaping. Sensitivity analyses show that whether permitting vaping is cost-effective or not depends on levels of vaping and smoking relapse in release; there is little evidence to inform these parameters.

## Declarations

### Ethics approval and consent participate

The overall study obtained ethical approval from the SPS Research Access and Ethics Committee. The health economic analysis also obtained approval from the University of Stirling, General Research Ethics Panel.

## Conflicts-ICJME

NM, AB, CB, ED, AHL, NL, JPP, SS, EJT, CWB, KH, KAB, have no conflicts of interest

LB is Chief Social Policy Adviser to the Scottish Government and Senior Responsible Officer for the place and wellbeing programme (population health). She took up this role, which is a part time secondment from Sept 2021 after the research was conducted.

## Contributor statements: suggest use CRedIT

NM: conceptualisation, methodology, formal analysis, funding acquisition, writing (original draft, review and editing) and approving final draft.

AB: conceptualisation, methodology, funding acquisition, writing (review and editing) and approving final draft

CB: conceptualisation, writing (review and editing) and approving final draft

ED: conceptualisation, methodology, writing (review and editing) and approving final draft AHL: methodology, writing (review and editing) and approving final draft

LB: conceptualisation, methodology, funding acquisition and approving final draft NL: design, reviewing and editing, and approving final draft

JPP: conceptualisation, funding acquisition and approving final draft

SS: conceptualisation, methodology, funding acquisition, writing (review and editing) and approving final draft.

EJT: conceptualisation, approving final draft

CW-B: writing (review and editing) and approving final draft CWB: conceptualisation and approving final draft

KH: conceptualisation, methodology, funding acquisition, writing (review and editing) and approving final draft

KB: conceptualisation, methodology, funding acquisition, writing (review and editing) and approving final draft

## Data Availability

All data produced in the present study are available upon reasonable request to the authors

## References

1. Office for National Statistics. Adult smoking habits in the UK - Office for National Statistics: Annual Population Survey from the Office for National Statistics; 2024 [Available from: https://www.ons.gov.uk/peoplepopulationandcommunity/healthandsocialcare/healthandlifeexpectancies/bulletins/adultsmokinghabitsingreatbritain/2022.

2. Hiscock R, Bauld L, Amos A, Platt S. Smoking and socioeconomic status in England: the rise of the never smoker and the disadvantaged smoker. Journal of Public Health. 2012;34(3):390–6.

3. ASH. Health Inequalities 2024 [Available from: https://ash.org.uk/uploads/ASH-Briefing_Health-Inequalities.pdf?

4. The Scottish Centre for Crime and Justice. Scotland’s Prison Population 2019 [Available from: https://www.sccjr.ac.uk/wp-content/uploads/2019/10/7-Scotlands-prison-population.pdf.

5. Carnie J, Broderick R, Cameron J, Downie D, Williams G. Scottish Prison Service 16th Prisoner Survey 2017 [Available from: https://www.sps.gov.uk/Corporate/Publications/Publication-6101.aspx.

6. Demou E, Dobson R, Sweeting H, Brown A, Sidwell S, O’Donnell R, et al. From Smoking-Permitted to Smokefree Prisons: A 3-Year Evaluation of the Changes in Occupational Exposure to Second-Hand Smoke Across a National Prison System. Annals of Work Exposures and Health. 2020;64(9):959–69.

7. Perrett SE, Craddock C, Dunseath G, Shankar G, Luzio S, Gray BJ. Evaluating the impact of a prison smoking ban on the cardiovascular health of men in a UK prison. International Journal of Prisoner Health. 2023;19(3):340–9.

8. Tweed E, Mackay D, Boyd K, Brown A, Byrne T, Conaglen P, et al. Evaluating the health impacts of a national smokefree prisons policy using trends in medication dispensing: an interrupted time series analysis as part of the Tobacco InPrisons study (TIPs). The Lancet Public Health. 2021.

9. Binswanger IA, Carson EA, Krueger PM, Mueller SR, Steiner JF, Sabol WJ. Prison tobacco control policies and deaths from smoking in United States prisons: population based retrospective analysis. Bmj-British Medical Journal. 2014;349.

10. Carnie J, Broderick R. Scottish Prison Service 17th Prisoner Survey 2019 [Available from: https://www.sps.gov.uk/sites/default/files/2024-02/17thPrisonSurvey_2019_Research.pdf.

11. National Institute for Health and Care Excellence. Tobacco: preventing uptake, promoting quitting and treating dependence, NICE guidance NG209 2023 [Available from: https://www.nice.org.uk/guidance/ng209.

12. Tang M-S, Tang Y-L. Can electronic-cigarette vaping cause cancer? Journal of cancer biology. 2021;2(3):68–70.

13. Martheswaran T, Shmunes MH, Ronquillo YC, Moshirfar M. The impact of vaping on ocular health: a literature review. International Ophthalmology. 2021;41(8):2925–32.

14. Bush A, Lintowska A, Mazur A, Hadjipanayis A, Grossman Z, del Torso S, et al. E-Cigarettes as a Growing Threat for Children and Adolescents: Position Statement From the European Academy of Paediatrics. Frontiers in Pediatrics. 2021;9.

15. Brown A, Woods-Brown C, Angus K, McMeekin N, Hunt K, Demou E. Recent evidence on rates and factors influencing smoking behaviours after release from smokefree prisons: a scoping review. Accepted in International Journal of Prison Health. 2024.

16. Caleyachetty A, Lewis S, McNeill A, Leonardi-Bee J. Struggling to make ends meet: exploring pathways to understand why smokers in financial difficulties are less likely to quit successfully. European Journal of Public Health. 2012;22:41–8.

17. Outside. F. 27,000 children in Scotland are given new rights: @familiesoutside; 2024 [Available from: https://www.familiesoutside.org.uk/27000-children-in-scotland-are-given-new-rights/.

18. Svanes C, Omenaas E, Jarvis D, Chinn S, Gulsvik A, Burney P. Parental smoking in childhood and adult obstructive lung disease: results from the European Community Respiratory Health Survey. Thorax. 2004;59(4):295–302.

19. West HW, Juonala M, Gall SL, Kahonen M, Laitinen T, Taittonen L, et al. Exposure to Parental Smoking in Childhood Is Associated With Increased Risk of Carotid Atherosclerotic Plaque in Adulthood The Cardiovascular Risk in Young Finns Study. Circulation. 2015;131(14):1239–46.

20. Leonardi-Bee J, Jere ML, Britton J. Exposure to parental and sibling smoking and the risk of smoking uptake in childhood and adolescence: a systematic review and meta-analysis. Thorax. 2011;66(10):847–55.

21. Belvin C, Britton J, Holmes J, Langley T. Parental smoking and child poverty in the UK: an analysis of national survey data. Bmc Public Health. 2015;15.

22. Taking Action on Smoking and Health, ASH. Cost of living, cost of smoking: The effect of smoking on personal finances and poverty in Scotland 2023 [Available from: https://ashscotland.org.uk/wp-content/uploads/2024/06/The-Effect-of-Smoking-on-Personal-Finances-and-Poverty-in-Scotland-report-FINAL_12_2023.pdf.

23. Families Outside. Paying the Price. 2023.

24. Hunt KB, A. Eadie, D. McMeekin, N. Boyd, K. Bauld, L., et al. Process and impact of implementing a smoke-free policy in prisons in Scotland: TIPs mixed-methods study. Public Health Research. 2022;10(1).

25. McMeekin N, Wu O, Boyd KA, Brown A, Tweed EJ, Best C, et al. Implementation of a national smoke-free prison policy: an economic evaluation within the Tobacco in Prisons (TIPs) study. Tobacco Control. 2023;32(6):701–8.

26. National Institute for Health and Care Excellence. Methods for the development of NICE public health guidance | 6 Incorporating health economics 2012 [Third edition:[Available from: https://www.nice.org.uk/process/pmg4/chapter/incorporating-health-economics.

27. Husereau D, Drummond M, Augustovski F, De Bekker-Grob E, Briggs AH, Carswell C, et al. Consolidated health economic evaluation reporting standards 2022 (CHEERS 2022) statement: updated reporting guidance for health economic evaluations. International Journal of Technology Assessment in Health Care. 2022;38(1).

28. Jin X, Kinner SA, Hopkins R, Stockings E, Courtney RJ, Shakeshaft A, et al. Brief intervention on Smoking, Nutrition, Alcohol and Physical (SNAP) inactivity for smoking relapse prevention after release from smoke-free prisons: a study protocol for a multicentre, investigator-blinded, randomised controlled trial. BMJ open. 2018;8(10):e021326.

29. Scottish Government. The Scottish Health Survey 2022 [Available from: https://www.gov.scot/publications/scottish-health-survey-2022-volume-1-main-report/pages/11/.

30. Jones M, Smith M, Lewis S, Parrott S, Coleman T. A dynamic, modifiable model for estimating cost-effectiveness of smoking cessation interventions in pregnancy: application to an RCT of self-help delivered by text message. Addiction. 2019;114(2):353–65.

31. Egger S, Watts C, Dessaix A, Brooks A, Jenkinson E, Grogan P, et al. Parent’s awareness of, and influence on, their 14-17-year-old child’s vaping and smoking behaviours; an analysis of 3242 parent-child pairs in Australia. Addictive Behaviors. 2024;150.

32. National Center for Chronic Disease Prevention and Health Promotion (US) Office on Smoking and Health. The Health Consequences of Smoking - 50 Years of Progress: A Report of the Surgeon General. 2014.

33. Chaffee BW, Barrington-Trimis J, Liu F, Wu R, McConnell R, Krishnan-Sarin S, et al. E-cigarette use and adverse respiratory symptoms among adolescents and Young adults in the United States. Preventive Medicine. 2021;153.

34. Tackett AP, Urman R, Barrington-Trimis J, Liu F, Hong H, Pentz MA, et al. Prospective study of e-cigarette use and respiratory symptoms in adolescents and young adults. Thorax. 2024;79(2):163–8.

35. Oberg M, Jaakkola MS, Woodward A, Peruga A, Pruess-Ustuen A. Worldwide burden of disease from exposure to second-hand smoke: a retrospective analysis of data from 192 countries. Lancet. 2011;377(9760):139-46.

36. National Records of Scotland. Statistics and Data, Vital Events, Table 5.01(b) 2022 [Available from: https://www.nrscotland.gov.uk/statistics-and-data/statistics/statistics-by-theme/vital-events/general-publications/vital-events-reference-tables/2022/list-of-data-tables#section5.

37. Doll R, Peto R, Wheatley K, Gray R, Sutherland I. MORTALITY IN RELATION TO SMOKING - 40 YEARS OBSERVATIONS ON MALE BRITISH DOCTORS. British Medical Journal. 1994;309(6959):901-11.

38. Graham L, Fischbacher CM, Stockton D, Fraser A, Fleming M, Greig K. Understanding extreme mortality among prisoners: a national cohort study in Scotland using data linkage. European Journal of Public Health. 2015;25(5):879–85.

39. Spaulding AC, Eldridge GD, Chico CE, Morisseau N, Drobeniuc A, Fils-Aime R, et al. Smoking in Correctional Settings Worldwide: Prevalence, Bans, and Interventions. Epidemiologic Reviews. 2018;40(1):82–95.

40. Zlodre J, Fazel S. All-cause and external mortality in released prisoners: systematic review and meta-analysis. American journal of public health. 2012;102(12):e67–75.

41. Parrott S, Godfrey C, Raw M, West R, McNeill A. Guidance for commissioners on the cost effectiveness of smoking cessation interventions. Thorax. 1998;53:AS1-AS38.

42. Asthma and Lung UK. Investing in breath: Measuring the economic cost of asthma and COPD in the UK and identifying ways to reduce it through better diagnosis and care 2023 [Available from: https://www.asthmaandlung.org.uk/investing-breath-measuring-economic-cost-asthma-copd-uk-identifying-ways-reduce-it-through-better.

43. Jones M, et al. Economics of Smoking in Pregnancy (ESIP) Model - The University of Nottingham 2019 [Available from: https://www.nottingham.ac.uk/research/groups/tobaccoandalcohol/smoking-in-pregnancy/esip/index.aspx.

44. Cancer Research UK. Is vaping harmful? 2022 [updated 2018-12-28. Available from: https://www.cancerresearchuk.org/about-cancer/causes-of-cancer/smoking-and-cancer/is-vaping-harmful.

45. Jones KC, Weatherly H, Birch S, Castelli A, Chalkley M, Dargan A, et al. Unit Costs of Health and Social Care 2022 Manual. Personal Social Services Research Unit (University of Kent) & Centre for Health Economics (University of York), Kent, UK 2023.

46. McNamara S, Schneider PP, Love-Koh J, Doran T, Gutacker N. Quality-Adjusted Life Expectancy Norms for the English Population. Value in Health. 2023;26(2):163–9.

47. Maheswaran H, Petrou S, Rees K, Stranges S. Estimating EQ-5D utility values for major health behavioural risk factors in England. Journal of Epidemiology and Community Health. 2013;67(2):172–80.

48. Li J, Hajek P, Pesola F, Wu Q, Phillips-Waller A, Przulj D, et al. Cost-effectiveness of e-cigarettes compared with nicotine replacement therapy in stop smoking services in England (TEC study): a randomized controlled trial. Addiction. 2020;115(3):507–17.

49. Sullivan PW, Slejko JF, Sculpher MJ, Ghushchyan V. Catalogue of EQ-5D Scores for the United Kingdom. Medical Decision Making. 2011;31(6):800–4.

50. Briggs A, Claxton K, Sculpher M. Decision Modelling for Health Economic Evaluation. 2011 ed. Gray A, Briggs A, editors. New York: Oxford University Press; 2006.

51. Li L, Nguyen C, Lin Y, Guo Y, Abou Fadel N, Zhu Y. Impacts of electronic cigarettes usage on air quality of vape shops and their nearby areas. Science of the Total Environment. 2021;760.

52. Protano C, Manigrasso M, Cammalleri V, Zoccai GB, Frati G, Avino P, et al. Impact of Electronic Alternatives to Tobacco Cigarettes on Indoor Air Particular Matter Levels. International Journal of Environmental Research and Public Health. 2020;17(8).

53. Brown A, O’Donnell R, Eadie D, Purves R, Sweeting H, Ford A, et al. Initial Views and Experiences of Vaping in Prisons: A Qualitative Study With People in Custody Preparing for the Imminent Implementation of Scotland’s Prison Smokefree Policy. Nicotine & Tobacco Research. 2021;23(3):543–9.

54. Brown A, O’Donnell R, Eadie D, Ford A, Mitchell D, Hackett A, et al. E-cigarette Use in Prisons With Recently Established Smokefree Policies: A Qualitative Interview Study With People in Custody in Scotland. Nicotine & tobacco research: official journal of the Society for Research on Nicotine and Tobacco. 2021;23(6):939–46.

55. Jin X, Kinner SA, Hopkins R, Stockings E, Courtney RJ, Shakeshaft A, et al. A randomised controlled trial of motivational interview for relapse prevention after release from smoke-free prisons in Australia. International Journal of Prisoner Health. 2021;17(4):462–76.

56. Martin RA, Stein LAR, Kang A, Rohsenow DJ, Bock B, Martin SA, et al. Circumstances Around Cigarette Use after Enforced Abstinence From Smoking in an American Prison. Journal of Addiction Medicine. 2022;16(6):E395–E401.

57. Winkelman TNA, Ford BR, Dunsiger S, Chrastek M, Cameron S, Strother E, et al. Feasibility and Acceptability of a Smoking Cessation Program for Individuals Released From an Urban, Pretrial Jail A Pilot Randomized Clinical Trial. Jama Network Open. 2021;4(7).

58. Scottish Government. Reconviction Rates in Scotland: 2019-2020 Offender Cohort. 2023.

59. UK Government. Advancing our health: prevention in the 2020s – consultation document. 2019.

60. Khan J. The Khan Review: Making Smoking Obsolete. 2022.

61. Zhang CX, Lewer D, Aldridge RW, Hayward AC, Cornaglia C, Trussell P, et al. Small numbers, big impact: making a utilitarian case for the contribution of inclusion health to population health in England. Journal of Epidemiology and Community Health. 2023;77(12):816–20.

62. Puljevic C, de Andrade D, Coomber R, Kinner SA. Relapse to smoking following release from smoke-free correctional facilities in Queensland, Australia. Drug and Alcohol Dependence. 2018;187:127–33.

63. Albany H, Richmond R, Simpson M, Kariminia A, Hwang YI, Butler T. Smoking Beyond Prison Bans: The Impact of Prison Tobacco Bans on Smoking Among Prison Entrants. Journal of Correctional Health Care. 2021;27(4):280–8.

64. National Institute for Health and Care Excellence. Overview | Chronic obstructive pulmonary disease (acute exacerbation): antimicrobial prescribing | Guidance | NICE: NICE; 2024 [Available from: https://www.nice.org.uk/guidance/ng114.

65. Scottish Prison Service. Scottish Prison Population Statistics 2022-23 2023 [Available from: https://www.gov.scot/binaries/content/documents/govscot/publications/statistics/2023/11/scottish-prison-population-statistics-2022-23/documents/report-22-23/report-22-23/govscot%3Adocument/Scottish%2BPrison%2BPopulation%2BStatistics%2B2022-23%2B-%2BAnalyt#:~:text=The%20overall%20prison%20population%20remained,in%202022%2D23%20was%207%2C426.

