## Supplementary material for "How can we maximise the benefits of smoke-free prisons? Decision analytic model to predict potential impacts on public health"

### SUPPLEMENTARY MATERIALS

*Table S1 Model input parameters*

| Parameter | Basecase value | Source | PSA distributions (alpha, beta) | Sensitivity analysis value |
|---|---|---|---|---|
| **Transition probabilities** | | | | |
| <u>Smoking status:</u> | | | | |
| Prevalence vaping in prison (relevant to vaping permitted comparators) | 76% | SPS canteen data (Cath Best analysis) | N/A | 60% SPS 17th prisoner survey (2019) (1) |
| Tobacco smoking prevalence pre-prison (relevant to vaping not permitted comparators) | 68% | SPS 16th prisoner survey (2017) (2) | N/A | N/A |
| Smoking status on release – vaping permitted comparators | 92% relapse to smoking (therefore assume 8% will continue vaping) | Jin et al 2021 (3) RCT - figure comes from control arm | N/A | 1) 100% relapse to smoking (0% continue to vape)<br>2) 50% relapse to smoking (50% continue to vape) |
| Smoking status on release - no vaping permitted comparators | 92% smokers (8% non- | Jin et al 2021 (3) | N/A | 1) 94% Puljevic et al. (4) (6% non-smokers/non-vapers ) |

| Parameter | Basecase value | Source | PSA distributions (alpha, beta) | Sensitivity analysis value |
|---|---|---|---|---|
| | smokers/non-vapers) | RCT - figure comes from control arm | | 2) 86% Albany et al. (5) (14% non-smokers/non-vapers ) |
| Partner smoking status | Non-smoker/non vaper: 63%<br>Smoker: 21%<br>Former smoker: 6%<br>Vaper: 10% | Scottish Health Survey (2022) (6) | Beta (34.91, 20.5)<br>Beta (75.662, 284.632)<br>N/A<br>Beta (86.34, 777.02) | N/A |
| *Morbidity:* | | | | |
| Vapers lung injury (bronchitis) | 4.9% | Chaffee et al 2021 (7) | Beta (17.278, 335.33) | 1) 9.2%<br>2) 4.7% |
| Smoking related disease prevalence (COPD, CHD, lung cancer and stroke) | Various (age and sex dependent) | Jones et al 2018 (8) | N/A | N/A |
| Relative risks for smoking related diseases (COPD, CHD, lung cancer and stroke) | Various (age and smoking status dependent) | National Center for Chronic Disease Prevention and Health Promotion (US) Office on Smoking and Health 2014 (9) | Log normal (various) | N/A |
| Exposure to second-hand smoke (CHD and lung disease) | Various | Oberg et al (2011) (10) | Log normal (various) | N/A |
| *Mortality:* | | | | |
| Non-smoker and non-vaper | Various (age and sex dependent) | National records of Scotland (2022) (11) | N/A | N/A |
| Relative risk due to smoking status (former and current) | Various (age and smoking | Doll et al (1994) & Scottish Health Survey (6, 12) | Log normal (various) | N/A |

| Parameter | Basecase value | Source | PSA distributions (alpha, beta) | Sensitivity analysis value |
|---|---|---|---|---|
| | status dependent) | | | |
| Exposure to second-hand smoke (CHD and lung disease) | 1% | Oberg et al (2011) (10) | N/A | |
| **Health related utilities** | | | | |
| Non-smoker and ex-vaper | Various (age and sex dependent) | McNamara et al (2022) (13) | N/A | N/A |
| Non-smoker/non-vaper exposed to SHS (disutility) | -0.02 | McMeekin et al. (14) | Beta (94.10, 4610.9) | N/A |
| Tobacco smoker (disutility) | -0.062 | Maheswaran et al (2013) (15) | Beta (34.57, 522.96) | N/A |
| Former smoker (disutility) | -0.023 | Maheswaran et al (2013) (15) | Beta (27.46, 1166.3) | N/A |
| Smoking related disease (disutilities) | COPD: -0.1336<br>CHD: -0.0627<br>LC: -0.1192<br>Stroke -0.1171 | Sullivan et al (2011) (16) | Beta (30.96, 200.8)<br>Beta (21.41, 320.05)<br>Beta (6.65, 49.13)<br>Beta (82.57, 622.58) | N/A |
| Vaper (disutility) | -0.023 | Li et al (2020) (17) | N/A | N/A |
| Bronchitis | -0.0379 | NICE COPD NG114 (2018) (18) | Beta (6043.5, 153530) | N/A |
| **Costs** | | | | |
| E-cigarette spend | £600 annually | CRUK (2022) (19) | Gamma (88.51, 6.78) | N/A |
| Tobacco spend | £2,100 annually | CRUK (2022) (19) | Gamma (88.31,23.78) | N/A |
| Healthcare SRD | COPD: £930<br>CHD: £2,027<br>LC: £10,535<br>Stroke: £4,793 | Jones et al (2019) (8) | Gamma (99.99, 9.30)<br>Gamma (100, 20.27)<br>Gamma (100, 105.35)<br>Gamma (21583,0.22) | N/A |
| Vaping related lung disease | £257.08 | Asthma and lung UK (2023) (20) | N/A | N/A |
| **Intervention** | | | | |

| Parameter | Basecase value | Source | PSA distributions (alpha, beta) | Sensitivity analysis value |
|---|---|---|---|---|
| Intervention engaging | 53% tobacco smokers would like to give up smoking | 16th SPS prisoner survey (2) | N/A | 66% of smokers report wanting to give up 2022 SHS |
| Intervention effectiveness and cost | 6% quit smoking, cost £111.10 | NICE NG209 guideline (21) | | 1) 4% quit smoking, cost £10.67<br>2) 15% quit smoking, cost £122.96 |
| **Miscellaneous** | | | | |
| Child uptake of smoking/vaping – parent non-smoker | 0.152 | Jones et al (2019) (8) | Beta (369.79, 2067.6) | 1) 0.2275<br>2) 0.0076 |
| RR child uptake of vaping (compared to parent non-smoker) | Parent ever vaper: 1.42<br>Parent ever smoker: 1.81 | Egger et al (2024) (22) | N/A | N/A |
| RR child uptake of smoking (compared to parent non-smoker) | Parent ever vaper: 1.97<br>Parent ever smoker: 2.59 | Egger et al (2024) (22) | N/A | N/A |
| Age released prisoner | 37 | SPS population statistics (2022/23) (23) | N/A | N/A |
| Age - partner | 37 | Assumption | N/A | N/A |
| Age - child | 15 | Assumption | N/A | N/A |
| Discount rate | 1.5% | NICE (2012) (24) | N/A | N/A |

CHD–coronary heart disease; chronic obstructive pulmonary disease–COPD; LC–lung cancer; SHS–second-hand smoke); SRD–smoking related disease

*Table S2 Sensitivity analysis results - Released people*

| Sensitivity analysis | | Permitting vaping v. not permitting vaping with no intervention in smokefree prison (Difference 1) | | Vaping permitted in smokefree prison (Difference 2) | | No vaping permitted in smokefree prison (Difference 3) | |
|---|---|---|---|---|---|---|---|
| | | Vaping permitted (Scenario 1) | No vaping permitted (Scenario 2) | No intervention (Scenario 1) | Intervention (Scenario 3) | No vaping permitted (Scenario 2) | Intervention (Scenario 4) |
| | | Incremental | | | | | |
| | Basecase | Costs | QALYs | Costs | QALYs | Costs | QALYs |
| | | £3,764 (95% CI £2,715 to £5,125) | -0.252 (95% CI -0.355 to -0.17) | £1,022 (95% CI £866 to £1,209) | -0.044 (95% CI -0.056 to -0.033) | £704 (95% CI £561 to 871) | -0.037 (95% CI -0.126 to 0.045) |
| Vaping status in prison 60% | 76% | -£2,143 (95% CI -£3,323 to -£920) | 0.209 (95% CI 0.108 to 0.294) | £816 (95% CI £698 to £948) | -0.035 (95% CI -0.044 to -0.025) | £714 (95% CI £581 to £874) | -0.039 (95% CI -0.124 to 0.044) |
| Vaping permitted: 100% smoking on release | 92% | £5,226 (95% CI £3,898 to £6,737 | -0.416 (95% CI -0.534 to -0.32 | £865 (95% CI £697 to £1,063 | -0.046 (95% CI -0.057 to -0.034) | £712 (95% CI £569 to £879) | -0.039 (95% CI -0.131 to 0.045) |
| Vaping permitted: 50% smoking on release | | -£3,682 (95% CI -£5,936 to -£1,454) | 0.614 (95% CI 0.49 to 0.728) | £1,907(95% CI £1,754 to £2,076) | -0.036 (95% CI -0.047 to -0.024) | £708 (95% CI £562 to £867) | -0.041(95% CI -0.124 to 0.045) |

| | | | | | | | |
|---|---|---|---|---|---|---|---|
| No vaping permitted: 94% smoking on release (worst) | 92% | £3,229 (95% CI £2,299 to £4,658 | -0.208 (95% CI -0.315 to -0.135) | £1,023 (95% CI £862 to £1,201) | -0.044 (95% CI -0.055 to -0.032) | £720 (95% CI £570 to £878) | -0.039 (95% CI - 0.122 to 0.04) |
| No vaping permitted: 86% smoking on release (best) | | £5,374 (95% CI £4,177 to £6,870) | -0.377 (95% CI -0.477 to -0.29) | £1,029 (95% CI £874 to £1,200) | -0.044 (95% CI -0.055 to -0.031) | £664 (95% CI £526 to £806) | -0.036 (95% CI -0.117 to 0.047) |
| Vaping related lung disease 9.2% (worst) | 4.9% | £3,812 (95% CI £2,679 to £5,228) | -0.253 (95% CI -0.36 to - 0.169) | £1,029 (95% CI (£873 to £1,206) | -0.044 (95% CI -0.056 to -0.032) | £709 (95% CI £564 to £871) | -0.037 (95% CI -0.12 to 0.05) |
| Vaping related lung disease 4.7% (best) | | £3,781(95% CI £2,758 to £5,112) | -0.250 (95% CI -0.345 to -0.169) | £1,031(95% CI £880 to £1,201) | -0.044 (95% CI -0.056 to -0.032) | £712 (95% CI £576 to £869) | -0.036 (95% CI -0.119 to 0.048) |
| Intervention engaging 66% (best) | 53% | £3,761 (95% CI £2,740 to £5,044) | -0.249 (95% CI -0.341 to -0.173) | £1,225 (95% CI £1,019 to £1,439) | -0.055 (95% CI -0.069 to -0.039) | £882 (95% CI £700 to £1,078) | -0.046 (95% CI -0.128 to 0.037) |
| Intervention effectiveness and cost: low - 4% £10.67 | Mid - 6% and £111.10 | £3,766 (95% CI £2,718 to £5,188) | -0.248 (95% CI -0.354 to -0.171) | £784 (95% CI £681 to £897) | -0.030 (95% CI -0.038 to -0.021) | £495 (95% CI £401 to £596) | -0.026 (95% CI -0.103 to 0.057) |
| Intervention effectiveness: high – 15% £122.96 | | £3,770 (95% CI £2,721 to £5,177) | -0.250 (95% CI -0.351 to -0.17 | £2,309 (95% CI £1,921 to £2,765) | -0.109 (95% CI -0.138 to -0.08 | £1,825 (95% CI £1,463 to £2,234) | -0.095 (95% CI -0.178 to -0.002 |

*Table S3 Sensitivity analysis - Child*

| **Uptake of smoking in children living with parent who smokes (basecase 15.2%)** | **Living with nicotine-free released prisoner (Scenario 1)** | | **Living with released prisoner who smokes (Scenario 2)** | | **Living with released prisoner who vapes (Scenario 3)** | |
|---|---|---|---|---|---|---|
| | **Mean** | **95% CI** | **Mean** | **95% CI** | **Mean** | **95% CI** |
| **Worst (22.8%)** | | | | | | |
| **Total costs** | £18,623 | £ 15,970 to £21,520 | £41,777 | £35,092 to £48,988 | £32,480 | £27,398 to £37,979 |
| **Life-years** | 25.62 | 25.58 to 25.65 | 25.32 | 25.23 to 25.40 | 25.44 | 25.37 to 25.50 |
| **QALYs** | 21.81 | 21.66 to 21.94 | 20.90 | 20.59 to 21.17 | 21.26 | 21.02 to 21.48 |
| **Best (7.6%)** | | | | | | |
| **Total costs** | £7,725 | £6,521 to £8,999 | £15,444 | £12,420 to £18,601 | £12,345 | £10,034 to £14,771 |
| **Life-years** | 25.75 | 25.73 to 25.76 | 25.65 | 25.60 to 25.68 | 25.69 | 25.65 to 25.71 |
| **QALYs** | 22.23 | 22.12 to 22.32 | 21.92 | 21.76 to 22.06 | 22.04 | 21.91 to 22.16 |
| **<u>Prevalence</u>** | | | | | | |
| **Basecase** | | | | | | |
| Non-nicotine | 70% | | 33% | | 49% | |

| | | | | | |
|---|---|---|---|---|---|
| Non-nicotine (exposed to SHS | 0% | | 0% | | 0% |
| Person who smokes | 15% | | 39% | | 30% |
| Person who used to smoke | 0% | | 0% | | 0% |
| Person who vapes | 15% | | 27% | | 22% |
| Person who used to vape | 0% | | 0% | | 0% |
| **Worst** | | | | | |
| Non-nicotine | 54% | | 0% | | 23% |
| Non-nicotine (exposed to SHS | 0% | | 0% | | 0% |
| Person who smokes | 23% | | 59% | | 45% |
| Person who used to smoke | 0% | | 0% | | 0% |
| Person who vapes | 23% | | 41% | | 32% |
| Person who used to vape | 0% | | 0% | | 0% |
| **Best** | | | | | |
| Non-nicotine | 85% | | 67% | | 74% |
| Non-nicotine (exposed to SHS | 0% | | 0% | | 0% |
| Person who smokes | 8% | | 20% | | 15% |
| Person who used to smoke | 0% | | 0% | | 0% |
| Person who vapes | 8% | | 14% | | 11% |
| Person who used to vape | 0% | | 0% | | 0% |
